# Seroprevalence of teratogenic viruses among women of childbearing age in Botswana

**DOI:** 10.1101/2024.09.20.24314038

**Authors:** I. Gobe, K. Baipoledi, G. Skwenje, M. Ntamo, M. Motswaledi

## Abstract

**Background:** Teratogenic viruses are viruses than can cross the placenta and infect a growing foetus, resulting in malformations and birth defects. Some of the commonly known teratogenic viruses include cytomegalovirus (CMV), rubella, herpes simplex and varicella zoster (VZV) viruses. Most birth defects associated with these infections affect the central nervous system and sensory organs leading to symptoms that include mental retardation, hearing loss and blindness. The economic burden caused by congenital birth defects is high, as many affected children require special care, therapeutic and educational services. Despite the risk posed by teratogenic viruses during pregnancy, there is no national screening for active CMV, Rubella or VZV infection during pregnancy in Botswana and most African countries. Furthermore, data on the seroprevalence of these viruses among women of childbearing age is limited.

**Methods and setting:** This cross-section study used eighty-nine (89) residual plasma samples from Scottish Livingstone Hospital Laboratory in Molepolole-Botswana. Samples were from women between the ages 15-49 years. Samples were tested for antibodies against rubella, VZV and CMV using enzyme linked immunosorbent assay.

**Results:** Our results show a high seroprevalence of rubella IgG antibodies (97%), even though a small proportion (3%) of women are still susceptible. There was also a high seroprevalence of CMV IgG (100%) which was accompanied by an equally high CMV IgM of 98%. Seroprevalence of VZV IgG was low (63%) and 3% of the samples showed active VZV infection.

**Conclusions:** Teratogenic viruses are a concern in the population. This calls for preventative measures which include prompt screening and vaccination of susceptible eligible women to prevent congenital abnormalities in children.

## Introduction

Teratogenic agents can interfere with normal fetal development. These agents include but not limited to drugs, viral infections, maternal conditions (e.g. diabetes, epilepsy) and radiation (Kaleelullah and Garugula, 2021). Viruses such as rubella virus, Cytomegalo virus (CMV) and varicella zoster virus (VZV) can cross the placenta and result in fetal tissue damage (Kaleelullah and Garugula, 2021).

CMV and VZV are double-stranded DNA viruses from the Herpesviridae family. Upon initial infection with one of these viruses, the virus persists indefinitely as a latent infection which could reactivate under immunosuppression (Richman et al., 2002). CMV infections are generally asymptomatic in both children and adults (Johnson et al., 2012). Transmission occurs through contact with body fluids such as urine and saliva (Pesch et al., 2021). However, congenital CMV infection is a major cause of mental retardation, hearing loss and cerebral palsy (Johnson et al., 2012). Congenital CMV infections may occur following a primary infection or as a re-infection of an expectant mother. Primary infections occur in individuals with low antibody titres against the virus (Johnson et al., 2012). Primary CMV infection poses a 30-40% risk of fetal transmission while non-primary or reactivation poses over 4% risk (Coppola et al., 2019). Generally, poorer fetal outcomes are common if the pregnant woman is infected during early gestation (Pesch et al., 2021). In areas with high CMV seroprevalence, seronegative mothers have high rates of CMV acquisition with high placental transmission (Coppola et al., 2019). Global CMV seroprevalence is 83% (Zuhair et al., 2019). High prevalence of >90% is seen in WHO eastern Mediterranean regions (Zuhair et al., 2019) and the African region (Mhandire et al., 2019). According to the Centres for Disease Control and Prevention [CDC], approximately 1 in 200 babies are born with congenital CMV; 1 in 5 of these will develop birth defects, the most common of which is hearing loss which can be detected after birth or later in childhood (CDC,2024).

Varicella zoster infections are transmitted through the respiratory route. Primary varicella zoster infection causes chicken pox while secondary/reactivation causes shingles (Bhavsar and Mangat, 2021). Due to circulating maternal antibodies, reactivation of varicella zoster (Shingles) does not pose a risk to developing fetus. Primary infection with chicken pox poses a risk to the fetus and can cause congenital varicella syndrome (Bhavsar and Mangat, 2021). Congenital varicella zoster viral infection is common if primary infection occurs in the first half of pregnancy with greatest risk between 13-20 weeks of gestation (Bhavsar and Mangat, 2021). Furthermore, infants who get exposed to the virus 5 days before and 2 days after delivery are at an increased risk of experiencing severe disease with increased mortality (Singh et al., 2022). Congenital varicella syndrome consists of skin lesions, neurologic defects, eye disease and skeletal anomalies (Sauerbrei and Wutzler, 2000). Unlike CMV, varicella zoster primary infections can be prevented by vaccination. In fact, World Health Organisation (WHO) recommends vaccination for all children. However, many developing countries and some developed countries like the UK have not included this vaccine in their routine childhood immunization schedules (Singh et al., 2022). The seroprevalence of VZV in developed vaccinating countries is high with USA recording a seroprevalence of >99% in persons 30yrs and older (Kilgore et al., 2003), while Canada recorded 95% in people with HIV (Zou et al., 2022). In Africa where the vaccine is not routinely administered the seroprevalence is low; in Sudan the VZV seroprevalence lies at 50% (Adam et al., 2023), while Nigeria recorded 66% prevalence among children (Oripelaye et al., 2024).

Rubella is an RNA virus from the Rubi virus genus (Das and Kielian, 2021). Rubella infections spread through respiratory droplets. These infections are generally mild and are characterised by a fever and rash. Nonetheless, in pregnancy, rubella infection can cause miscarriages or congenital rubella syndrome (CRS), (Das and Kielian, 2021). According to the WHO, infection of a pregnant woman in the first trimester carries a 90% risk of fetal transmission and consequently CRS. CRS symptoms include hearing impairment, eye, and heart defects. Other CRS associated disabilities include autism, diabetes mellitus and thyroid dysfunction (WHO, 2024). CRS is common in populations where women of childbearing age are seronegative. According to a survey carried out on reports from 1963-2009, the seroprevalence of Rubella in Africa ranged from 68-98% (Goodson et al., 2011). No data from Botswana was available in this report. However, neighbouring South Africa recorded a seroprevalence of 60% (Goodson et al., 2011). The WHO report of 2020 estimated that about 100,000 children are affected by CRS annually worldwide (WHO, 2020 rubella vaccine position paper). The best preventative measure against CRS is vaccination. A single dose of rubella vaccine gives 95% immunity which is similar to that achieved by natural infection (WHO, 2024). Botswana introduced rubella vaccine in its childhood immunisation schedule in 2016 (MoH-Botswana). This should increase the seroprevalence of rubella in the long run. However, there is still limited data on current seroprevalence of rubella in women of childbearing age.

The economic burden caused by congenital birth defects is high, as many affected children require special care, therapeutic and educational services. However, despite the risk posed by teratogenic viruses during pregnancy, there is no national screening for active CMV, Rubella or VZV infection during pregnancy in Botswana. Furthermore, data on the seroprevalence of these viruses among women of childbearing age is limited. Therefore, the aim of this study is to determine the seroprevalence of these teratogenic viruses among women of childbearing age in Botswana.

## Materials and methods

### Study design and settings

The cross-sectional study was conducted between October and December 2023. Residual plasma samples were collected from Scottish Livingstone hospital laboratory in Molepolole, Botswana. Samples were stored at -20^°^C until further testing at the School of Allied Health Science Laboratory, University of Botswana.

### Study population and sampling strategy

Eighty-nine, residual (EDTA-anticoagulated, separated) blood samples were collected from women of childbearing age (15-49 Years) from Scottish Livingstone hospital laboratory in Molepolole, Botswana. Non-probability convenience sampling was carried out to include all females who are in the childbearing age group. Only samples that were stored in the refrigerator for not more than a week were selected.

The sample size was calculated using a single population proportion sample size formula by Daniel (1999). A seroprevalence of 95% was used for sample size calculation for CMV and rubella. These proportions were deduced from an epidemiology review article on CMV in Africa (Mhandire et al., 2019) and a rubella seroprevalence study that was carried out in Western Cape, South Africa (Corcoran and Hardie, 2005). For VSV, a seroprevalence of 50.4% was used for the calculations as it has been observed in data from Sudan (Adam et al., 2023). Calculated sample size was 73 for both rubella and CMV and 385 for VSV.

### Ethical considerations

Ethical clearance was obtained from University of Botswana Office of Research and Development and the Health Research and Development committee (HDRC) at the Ministry of Health, Botswana. Permission was sought from Scottish Livingstone Hospital for sample collection. The residual samples from Scottish Livingstone Hospital Laboratory were de-identified and given new random identities (“001-089”). Demographic data collected included age and sex only.

### Data collection

Commercial enzyme-linked immunosorbent assay (ELISA) kits were supplied by My BioSource (San Diego, USA), which included: Rubella virus IgG ELISA kit-MBS495665, Human VZV IgG ELISA kit-MBS7612746, VZV IgM ELISA kit -MBS7612747, Human CMV IgG ELISA kit-MBS57269157 and human CMV IgM ELISA kit-MBS7269158. The ELISA kits were used in accordance with the manufacturer’s instructions to measure the concentrations of IgG/IgM against the VZV, CMV and Rubella.

#### Rubella

Plasma samples were diluted 1:100 and then tested according to manufacturer’s instructions. A standard curve was prepared using a set of standards containing 0, 10, 50, 100 IU/mL. These standards were supposed to yield OD_620_ of <0.2, >0.2, >0.7 and >1.1, respectively. Using the standard curve and the linear equation calculated, the concentration of each sample was calculated and interpreted as either positive or negative accordingly: >15 IU/mL: positive, 10-15 IU/mL: Equivocal and <10 IU/mL: negative.

#### VZV

Samples and controls were run according to the manufacturer’s instructions. Interpretation of results depended on the results of the controls. For the controls to pass, mean OD_450_ of the negative control should be ≤0.1 and the positive control was ≥0.8. A cut-off value was calculated as the mean OD of negative control +0.10. Samples with absorbance values less than the cut-off Value were NON-REACTIVE and were considered NEGATIVE for VZV-IgG/IgM. Samples with absorbance values greater than the cut-off value were considered POSITIVE for VZV-IgG/IgM.

#### CMV

Both CMV IgG-Ab and IgM ELISA kits applied the competitive enzyme immunoassay technique, and the intensity of colour was inversely proportional to the amount of antibody measured. Interpretation of results was aided by the standards that were provided. Standards A-F with concentrations ranging from 0, 50, 100, 250, 500 and 1000 mg/mL respectively were run with samples. A standard curve was plotted relating the intensity of the colour (O.D_450_) to the concentration of standards. The CMV IgG/IgM-Ab concentration in each sample was then calculated from this standard curve. Any sample with an OD_450_ of more or equal to that of Standard A (0 mg/mL) was deemed negative while samples with OD_450_ of less than that of Standard A were deemed positive.

### Data analysis

The statistical significance level was set at p<0.05. All statistical analyses were performed using Microsoft Excel 365. Both categorical and quantitative data were used in the study. To determine the prevalence/frequencies, calculations were total number of samples that tested positive for IgG or IgM antibodies divided by the total number of samples tested multiplied by 100. Bar charts and box plots were generated using GraphPad Prism 10.3.1.

## Results

A total of 89 samples from women of childbearing age was analysed. Their age ranged from 15-44 years, with a mean age of 30.4 years.

Out of the 89 samples tested 2% (n=2/89) were seronegative for CMV IgM. 100% (n=89/89) were positive for CMV-IgG while 98% (87/89) were positive for CMV-IgM (Figure 1.0).

**Figure 1.0.**
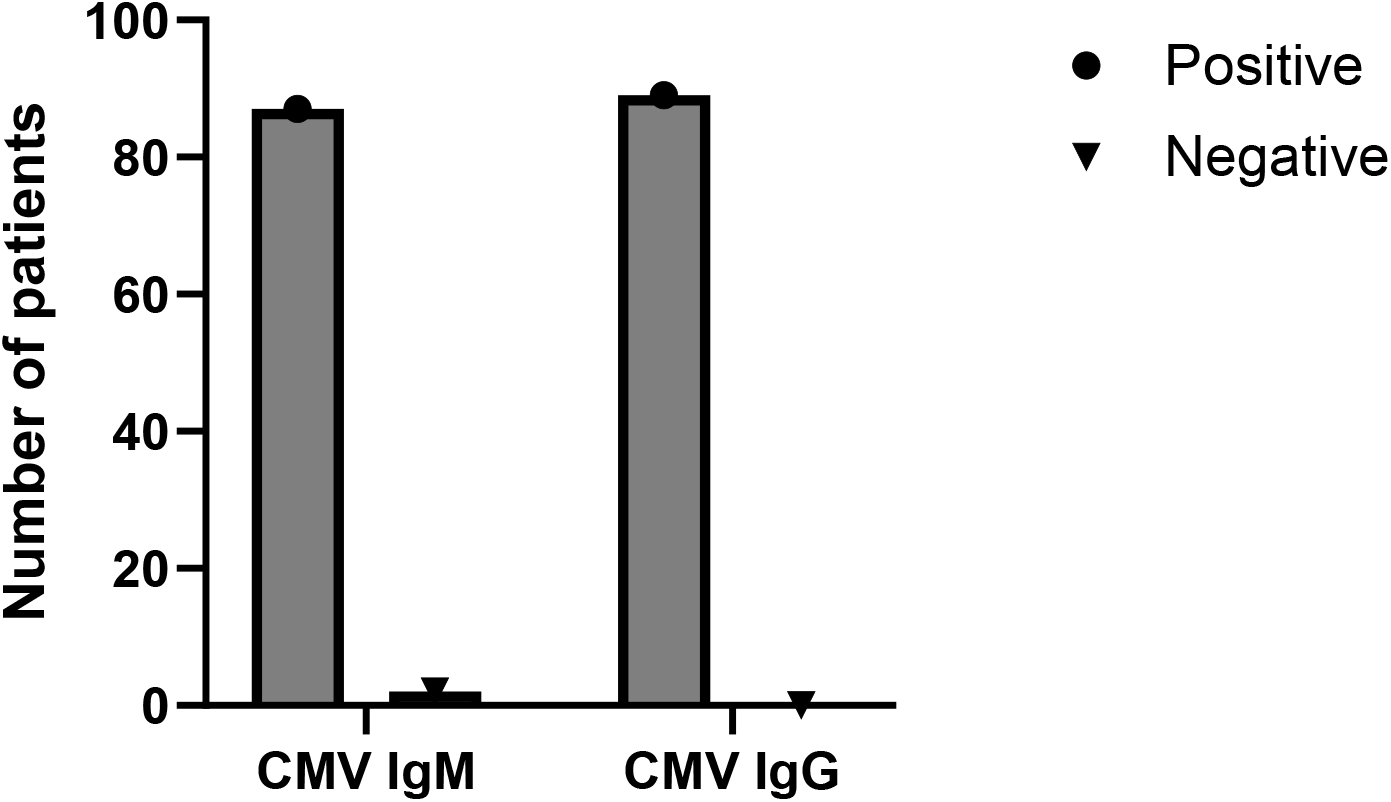
CMV IgM and IgG seroprevalence among 89 samples tested. Antibody testing was done using ELISA method.

Figure 2.0 below shows the concentration distribution of CMV-specific antibodies (IgG and IgM). The concentration of CMV IgG ranged from 32-476 mg/mL with a mean of 95.5 mg/mL with lower quartile and upper quartile of 65-95 mg/mL respectively. The concentration of CMV IgM ranged from 0-481 mg/mL with a mean of 209 mg/mL and lower quartile and upper quartile of 261 and 136 respectively. The mean concentration of IgM is higher than that of IgG with a statistically significant *p=* 0.005.

**Figure 2.0.**
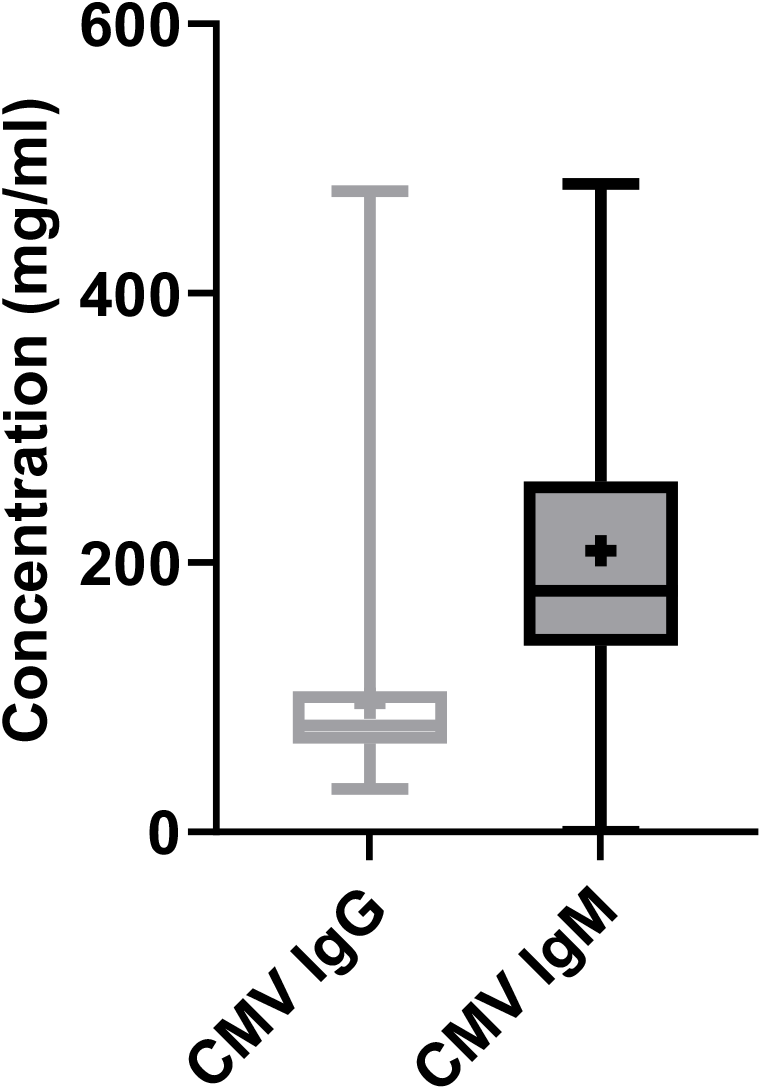
Concentration of CMV IgG and IgM in the sample population

Of the 89-plasma collected and tested for Rubella virus IgG antibodies, an overall seropositivity rate of 96.6 % (n=86/89) was observed. However, 3.4% (n=3/89) of the samples tested negative for Rubella virus IgG antibodies (Figure 3.0).

**Figure 3.0.**
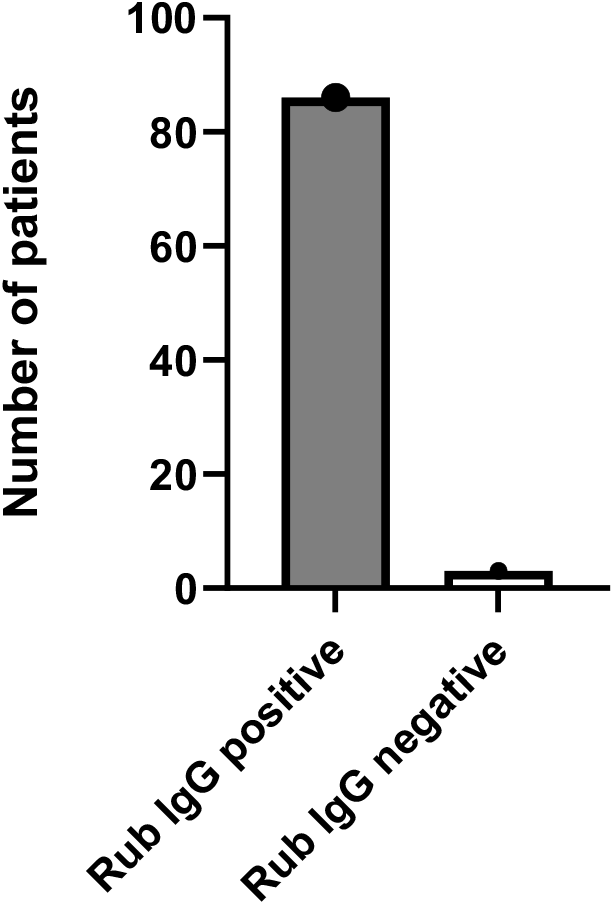
Rubella IgG seroprevalence among 89 samples tested. Antibody testing was done using ELISA method.

The concentration of Rubella IgG ranged from 1.1 -110 IU/mL with a mean of 62 IU/mL with a lower quartile and upper quartile of 50 and 75 IU/ML respectively (Figure 4.0).

**Figure 4.0.**
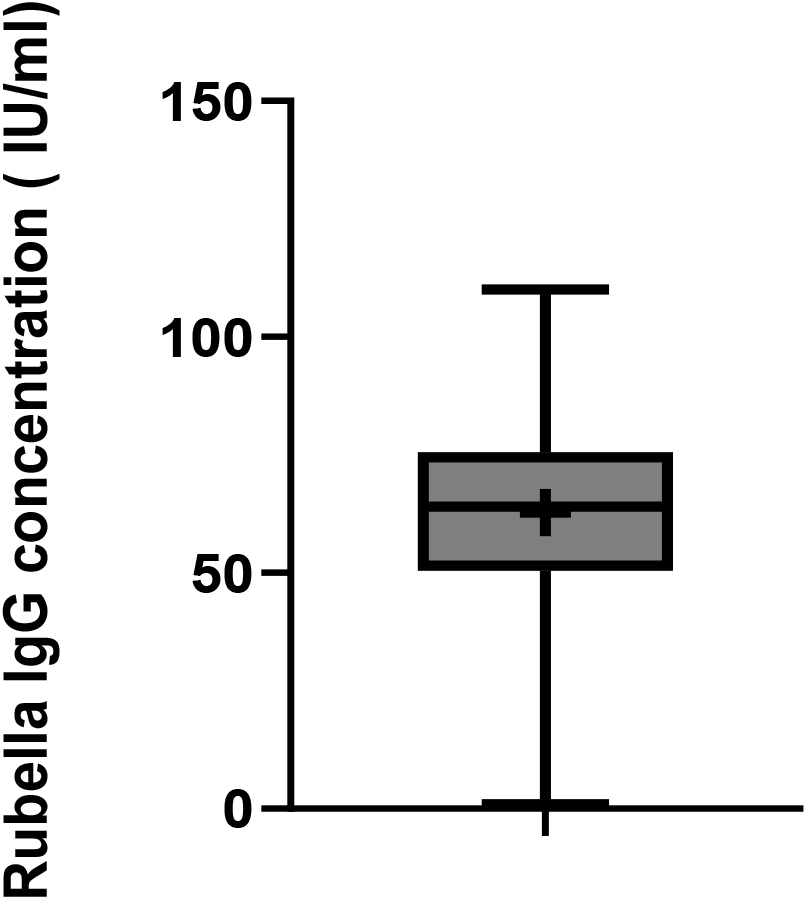
Concentration of Rubella IgG in the sample population

Figure 5.0 below shows the number of people who were seropositive for VZV-IgG and IgM. Out of the 89 samples, 53(63%) were positive for VZV IgG while 33 (37%) were negative for VZV IgG. 3% (n=3/89) were positive for VZV IgM and 86 (97%) were seropositive for VZV IgM. Among the three samples which were positive for IgM, 2 had both IgG and IgM while one was positive for IgM only.

**Figure 5.0.**
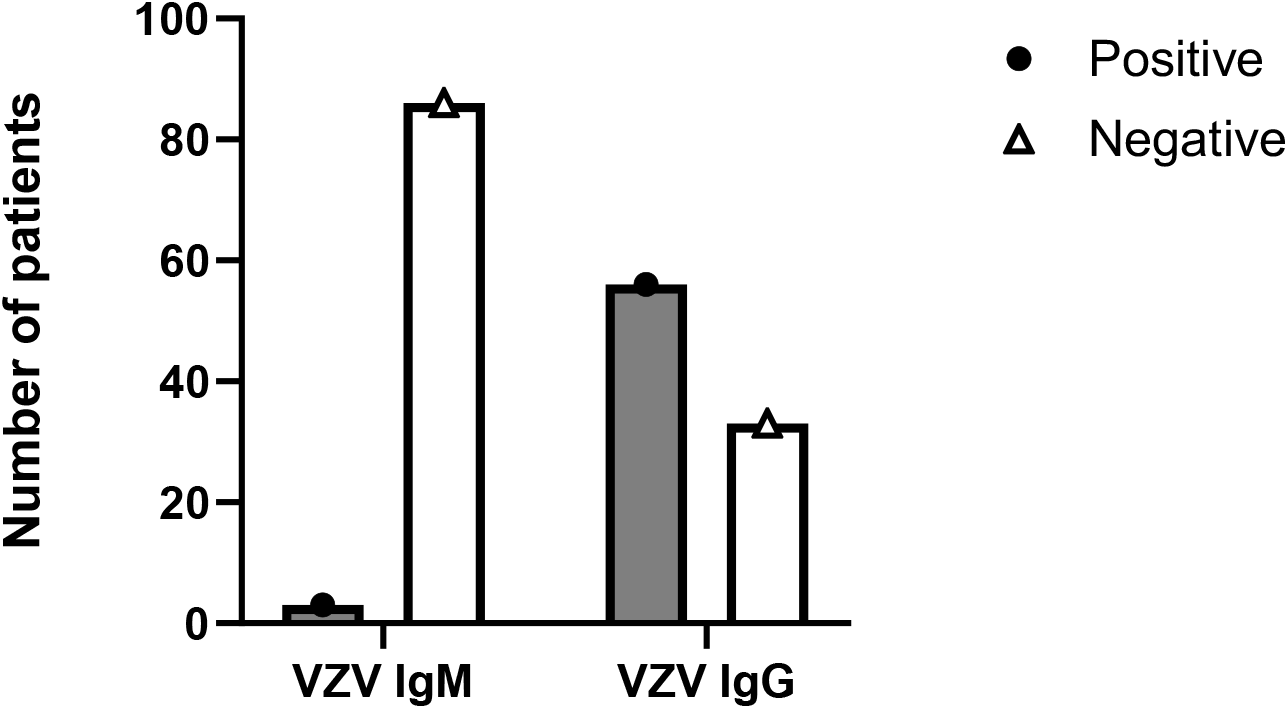
VZV IgG seroprevalence among 89 samples tested. Antibody testing was done using ELISA method.

## Discussion

### CMV

This study shows a high seroprevalence of CMV IgG which is 100%. The results are consistent with data from other African studies which estimate CMV seroprevalence among women to be >90% (Mhandire et al., 2019). Furthermore, the results are consistent with the global narrative that states that CMV seroprevalence correlates with socio-economic status and education levels, with high prevalence seen in under-developed and developing countries (Fowler et al., 2022). Our study also showed a high seroprevalence of CMV IgM in women of childbearing age, in comparison to the global seroprevalence of CMV IgM, which is estimated to be 13.7% (Adane and Getawa, 2021). These results were puzzling and alarming as they suggest a high risk for congenital malformations in Botswana. In Europe, North America and Latin America, the seroprevalence of CMV IgM is 1-4.6%, 2.3-4.5% and 0-0.7% respectively, among women (Fowler et al., 2022). In the African countries of Egypt, Kenya, Sudan, Ethiopia and Nigeria, the seroprevalence of CMV IgM was estimated to be 7.3%, 8.1%, 2.5%, 5.5% and 11.1% respectively (Hailemariam et al., 2021). Therefore, the CMV IgM seroprevalence found in this study is alarmingly high compared to other studies both in Europe and in Africa. CMV IgM is generally considered as a transient marker of primary infection. However, only 50% of individuals who are CMV IgM positive have actually been shown to have a primary infection (Prince and Lapé-Nixon, 2014). Studies have also shown that CMV IgM specificity is poor indicator of recent infection because it is produced during CMV viral reactivation, re-infection with a different CMV strains and it can also persist for over a year after primary CMV infection (Prince and Lapé-Nixon, 2014). The high seroprevalence of CMV IgM in our study could mean that active CMV infections are rife in our population or that IgM is not cleared after active infection. The latter might be true because in a study where CMV DNA was detected in HIV positive pregnant women in Botswana, the prevalence was found to be 14.6%(Moraka et al., 2019). Nonetheless, the high CMV IgM suggest something inherent in Botswana’s population that needs to be investigated further.

Since CMV IgM is inconclusive for primary infection and PCR testing is expensive, studies have shown that CMV IgG avidity testing is a more specific marker to identify pregnant women with recent CMV infection (Prince and Lapé-Nixon, 2014). Primary infections are worrisome because the risk of congenital CMV infection has been shown to be approximately 40% in infants born to mothers who acquire a primary CMV infection after conception (Prince and Lapé-Nixon, 2014). In spite of this, there is contradicting evidence that shows that in populations with very high CMV IgG seropositivity, such as Brazil (maternal CMV IgG seropositivity of >97%), there are more cases of congenital CMV infections (Mussi-Pinhata et al., 2018). This indicates that CMV IgG seropositivity is not necessarily protective. Despite high CMV seroprevalence in Africa, comparative data is lacking.

### Rubella

Our study shows that the seroprevalence of rubella is comparable to that found in other African countries which range from 68-98% (Goodson et al., 2011). A recent study from Morocco further recorded an IgG seroprevalence of 85% among pregnant women (Zahir et al., 2020). Generally, the cut-off point for protective rubella antibodies is 10 IUM/mL (Kempster et al., 2020). Our results show a mean protective immunity that is significantly higher than the recommended cut threshold. These results show that most of the women of childbearing age are protected from rubella infections. The Ministry of Health in Botswana introduced rubella vaccine in 2016, hence the vaccinated population are not yet at childbearing age. Therefore, protective immunity shown by this study reflects immunity due to natural infection. These results also show that rubella infections are very common which is a concern especially for those that are seronegative and are planning to be pregnant.

Although our study show that most participants are protected, there is still that 3% of women of childbearing age who still carry the risk of contracting rubella and consequently CRS, if they get pregnant. This calls for interventions such as screening and vaccinating those that are seronegative. Since rubella vaccine as a life attenuated vaccine may potentially be unsafe for pregnant women (WHO, 2024) it would be best to avail the vaccine to all women of childbearing age who are planning to be pregnant.

### VZV

Seroprevalence of VZV IgG among women of childbearing age in Botswana was the lowest with significant number of individuals being unprotected and susceptible. We found a small percentage of the study participants who had recent or active VSV infection as shown by positive IgM results in the absence of IgG. Limited data is available on the seroprevalence of VZV in Africa, the few studies done show a significantly low seroprevalence when compared to developing nations as found in Sudan which recorded a seroprevalence of 50% (Adam et al., 2023).The seroprevalence of VZV in developed vaccinating countries is high with USA recording a seroprevalence of >99% in persons 30 years and older (Kilgore et al., 2003), while Canada recorded 95% among people living with HIV (Zou et al., 2022). The low seroprevalence of VZV which indicates a high proportion of susceptible women is worrisome. Congenital VZV infections are common if primary infection occurs in the first half of pregnancy with greatest risk between 13-20 weeks of gestation (Bhavsar and Mangat, 2021). Furthermore, infants who get exposed to the virus 5 days before and 2 days after delivery are at an increased risk of experiencing severe disease with increased mortality (Singh et al., 2022). This calls for countries to heed WHO recommendation of including VZV vaccine in routine vaccination programs and target susceptible women who are planning to get pregnant.

## Limitations

There were several limitations to this study. First the sample size of the study was small. This was especially true for VZV data where an estimated sample size of 385 was supposed to be used but only 89 samples were analysed. Furthermore, co-morbidities which could affect the data such as HIV were not taken into consideration. Further studies need to be done to take into consideration these limitations.

## Conclusions

Our study shows a high seroprevalence of rubella IgG (97%), even though a small proportion (3%) of women are still susceptible. There is also a high seroprevalence of CMV IgG (100%). However, according to literature, this does not necessarily translate to low congenital CMV infections hence prompt screening of pregnant women for active CMV infections is necessary. The low seroprevalence of VZV IgG (63%) indicates that a significant proportion of the population is susceptible.

## Recommendations

The results call for improved strategies to reduce the burden of congenital defects. These could include early identification through maternal or new-born screening, vaccination, behavioural interventions and treatment for infected pregnant women and infants.

## Acknowledgements

The authors wish to express gratitude to the staff and management of Scottish Livingstone hospital laboratory in Molepolole, where samples were obtained.

## Competing interests

The authors declare that they have no financial or personal relationships that may have inappropriately influenced them in writing this article.

## Author contributions

Irene Gobe conceived the idea. The experiments were carried out by K. Baipoledi, G. Sekwenje and M. Ntamo under the supervision of Irene Gobe. I. Gobe also analysed the results and drafted the article. Prof. M. Motswaledi participated in data analysis, critiqued the content, and gave final approval of the version to be published.

## Data availability

Data sharing regarding this article is available upon written request to the corresponding author.

## Disclaimer

The views and opinions expressed in this article are those of the author(s) and do not necessarily reflect the official policy or position of any affiliated agency of the authors.

## Notes

### Competing Interest Statement

The authors have declared no competing interest.

### Funding Statement

This study was funded by Department of Tertiary Education Financing (DTEF)

### Author Declarations

Ethics committee/IRB of Ministry of Health Research and development division (MoH HRDC) gave ethical approval for this work. Also Ethics committee/IRB of University of Botswana gave ethical approval for this work.

## References

Adam, O., Musa, A., Kamer, A., Benharrat, S. & Hübschen, J. M. 2023. Active circulation of varicella zoster virus among different age groups in Sudan. Epidemiology & Infection, 151, e10.

Adane, T. & Getawa, S. 2021. Cytomegalovirus seroprevalence among blood donors: a systematic review and meta-analysis. Journal of International Medical Research, 49, 03000605211034656.

Bhavsar, S. M. & Mangat, C. 2021. Congenital varicella syndrome.

Coppola, T., Mangold, J. F., Cantrell, S. & Permar, S. R. 2019. Impact of maternal immunity on congenital cytomegalovirus birth prevalence and infant outcomes: a systematic review. Vaccines, 7, 129.

Corcoran, C. & Hardie, D. R. 2005. Seroprevalence of rubella antibodies among antenatal patients in the Western Cape. South African Medical Journal, 95.

Das, P. K. & Kielian, M. 2021. Molecular and structural insights into the life cycle of rubella virus. Journal of virology, 95, 10.1128/jvi. 02349–20.

Fowler, K., Mucha, J., Neumann, M., Lewandowski, W., Kaczanowska, M., Grys, M., Schmidt, E., Natenshon, A., Talarico, C. & Buck, P. O. 2022. A systematic literature review of the global seroprevalence of cytomegalovirus: possible implications for treatment, screening, and vaccine development. BMC Public Health, 22, 1659.

Goodson, J. L., Masresha, B., Dosseh, A., Byabamazima, C., Nshimirimana, D., Cochi, S. & Reef, S. 2011. Rubella epidemiology in Africa in the prevaccine era, 2002–2009. The Journal of infectious diseases, 204, S215–S225.

Hailemariam, M., Mekonnen, Z., Claeys, G. & Padalko, E. 2021. Congenital cytomegalovirus infections:(no) focus on Africa: a review. Gynecology and Obsteatrics, 11, 1–6.

Johnson, J., Anderson, B. & Pass, R. F. 2012. Prevention of maternal and congenital cytomegalovirus infection. Clinical obstetrics and gynecology, 55, 521–530.

Kaleelullah, R. A. & Garugula, N. 2021. Teratogenic genesis in fetal malformations. Cureus, 13.

Kempster, S. L., Almond, N., Dimech, W., Grangeot-KEROS, L., Huzly, D., Icenogle, J., El Mubarak, H. S., Mulders, M. N. & Nübling, C. M. 2020. WHO international standard for anti-rubella: learning from its application. The Lancet Infectious Diseases, 20, e17–e19.

Kilgore, P. E., Kruszon-Moran, D., Seward, J. F., Jumaan, A., Van Loon, F. P., Forghani, B., Mcquillan, G. M., Wharton, M., Fehrs, L. J. & Cossen, C. K. 2003. Varicella in Americans from NHANES III: implications for control through routine immunization. Journal of medical virology, 70, S111–S118.

Mhandire, D., Rowland-Jones, S., Mhandire, K., Kaba, M. & Dandara, C. 2019. Epidemiology of Cytomegalovirus among pregnant women in Africa. Journal of infection in developing countries, 13.

Moraka, N. O., Moyo, S., Mayondi, G., Leidner, J., Ibrahim, M., Smith, C., Weinberg, A., Li, S., Thami, P. K. & Kammerer, B. 2019. Cytomegalovirus Viremia in HIV-1 Subtype C Positive Women at Delivery in Botswana and Adverse Birth/Infant Health Outcomes. JAIDS Journal of Acquired Immune Deficiency Syndromes, 81, 118–124.

Mussi-Pinhata, M. M., Yamamoto, A. Y., Aragon, D. C., Duarte, G., Fowler, K. B., Boppana, S. & Britt, W. J. 2018. Seroconversion for cytomegalovirus infection during pregnancy and fetal infection in a highly seropositive population:”The BraCHS Study”. The Journal of infectious diseases, 218, 1200–1204.

Oripelaye, M. M., Olanrewaju, F. O., Ajani, A. A., Akinboro, A. O., Enitan, A. O. & Oninla, O. A. 2024. Seroprevalence of varicella-zoster virus among people living with human immunodeficiency virus in Ile-Ife, Nigeria: A cross-sectional study. Journal of Clinical Sciences, 21, 14–19.

Pesch, M. H., Kuboushek, K., Mckee, M. M., Thorne, M. C. & Weinberg, J. B. 2021. Congenital cytomegalovirus infection. BMJ, 373.

Prince, H. E. & LapÉ-Nixon, M. 2014. Role of cytomegalovirus (CMV) IgG avidity testing in diagnosing primary CMV infection during pregnancy. Clinical and Vaccine Immunology, 21, 1377–1384.

Sauerbrei, A. & Wutzler, P. 2000. The congenital varicella syndrome. Journal of Perinatology, 20, 548–554.

Singh, S., Sharma, A., Rahman, M. M., Kasniya, G., Maheshwari, A. & Boppana, S. B. 2022. Congenital and perinatal varicella infections. Newborn (Clarksville, Md.), 1, 278.

Zahir, H., Arsalane, L., Elghouat, G., Mouhib, H., Elkamouni, Y. & Zouhair, S. 2020. Seroprevalence of rubella in pregnant women in Southern Morocco. The Pan African Medical Journal, 35.

Zou, J., Krentz, H. B., Lang, R., Beckthold, B., Fonseca, K. & Gill, M. J. Seropositivity, risks, and morbidity from varicella-zoster virus infections in an adult PWH cohort from 2000–2020. Open Forum Infectious Diseases, 2022. Oxford University Press, ofac395.

Zuhair, M., Smit, G. S. A., Wallis, G., Jabbar, F., Smith, C., Devleesschauwer, B. & Griffiths, P. 2019. Estimation of the worldwide seroprevalence of cytomegalovirus: a systematic review and meta-analysis. Reviews in medical virology, 29, e2034.

Daniel W.W. (1999). Biostatistics: A foundation for Analysis in Health Sciences. 7th edition. New York: John Wiley & sons.

WHO, 2024. Rubella fact sheet. https://www.who.int/news-room/fact-sheets/detail/rubella. Retrieved 9/August/2024.

(WHO, 2024: https://www.who.int/news-room/fact-sheets/detail/rubella. Retrieved, 29 July 2024).

